# Seroneutralization of Omicron BA.1 and BA.2 in patients receiving anti-SARS-CoV-2 monoclonal antibodies

**DOI:** 10.1101/2022.03.09.22272066

**Authors:** Timothée Bruel, Jérôme Hadjadj, Piet Maes, Delphine Planas, Aymeric Seve, Isabelle Staropoli, Florence Guivel-Benhassine, Françoise Porrot, William-Henry Bolland, Yann Nguyen, Marion Casadevall, Caroline Charre, Hélène Péré, David Veyer, Matthieu Prot, Artem Baidaliuk, Lize Cuypers, Cyril Planchais, Hugo Mouquet, Guy Baele, Luc Mouthon, Laurent Hocqueloux, Etienne Simon-Loriere, Emmanuel André, Benjamin Terrier, Thierry Prazuck, Olivier Schwartz

**Affiliations:** Institut Pasteur, Université de Paris, CNRS UMR3569, Virus and Immunity Unit, 75015, Paris, France; Vaccine Research Institute, 94000, Créteil, France; Department of Internal Medicine, National Reference Center for Rare Systemic Autoimmune Diseases, AP-HP, APHP.CUP, Hôpital Cochin, Paris, France; KU Leuven, Department of Microbiology, Immunology and Transplantation, Laboratory of Clinical and Epidemiological Virology, Leuven, Belgium; CHR d’Orléans, service de maladies infectieuses, Orléans, France; Université de Paris, École doctorale BioSPC 562, 75013, Paris, France; Université de Paris, Faculté de Médecine, Paris, France; INSERM U1016, CNRS UMR8104, Institut Cochin, Paris, France; AP-HP, Laboratoire de Virologie, CHU Cochin, Paris, France; INSERM, Functional Genomics of Solid Tumors (FunGeST), Centre de Recherche des Cordeliers, Université de Paris and Sorbonne Université, Paris, France; Laboratoire de Virologie, Service de Microbiologie, Hôpital Européen Georges Pompidou, Assistance Publique des Hôpitaux de Paris, Paris, France; G5 Evolutionary Genomics of RNA Viruses, Institut Pasteur, Université de Paris, Paris, France; University Hospitals Leuven, Department of Laboratory Medicine, National Reference Centre for Respiratory Pathogens, Leuven, Belgium; Humoral Immunology Laboratory, Institut Pasteur, Université de Paris, INSERM U1222, Paris, France; KU Leuven, Department of Microbiology, Immunology and Transplantation, Laboratory of Clinical Microbiology, Leuven, Belgium

## Abstract

The SARS-CoV-2 Omicron BA.1 variant has been supplanted in many countries by the BA.2 sub-lineage. BA.2 differs from BA.1 by about 21 mutations in its spike. Human anti-spike monoclonal antibodies (mAbs) are used for prevention or treatment of COVID-19. However, the capacity of therapeutic mAbs to neutralize BA.1 and BA.2 remains poorly characterized. Here, we first compared the sensitivity of BA.1 and BA.2 to neutralization by 9 therapeutic mAbs. In contrast to BA.1, BA.2 was sensitive to Cilgavimab, partly inhibited by Imdevimab and resistant to Adintrevimab and Sotrovimab. Two combinations of mAbs, Ronapreve (Casirivimab + Imdevimab) and Evusheld (Cilgavimab + Tixagevimab), are indicated as a pre-exposure prophylaxis in immunocompromised persons at risk of severe disease. We analyzed sera from 29 such individuals, up to one month after administration of Ronapreve and/or Evusheld. After treatment, all individuals displayed elevated antibody levels in their sera and neutralized Delta with high titers. Ronapreve recipients did not neutralize BA.1 and weakly impaired BA.2. With Evusheld, neutralization of BA.1 and BA.2 was detected in 19 and 29 out of 29 patients, respectively. As compared to Delta, titers were more severely decreased against BA.1 (344-fold) than BA.2 (9-fold). We further report 4 breakthrough Omicron infections among the 29 participants. Therefore, BA.1 and BA.2 exhibit noticeable differences in their sensitivity to therapeutic mAbs. Anti-Omicron activity of Ronapreve, and to a lesser extent that of Evusheld, is reduced in patients’ sera, a phenomenon associated with decreased clinical efficacy.

## Introduction

The SARS-CoV-2 Omicron variant comprises three main sub-lineages termed BA.1, BA.2 and BA.3 (Viana et al., 2022). The original BA.1 sub-lineage (also termed B.1.1.529) was identified in November 2021 and became dominant worldwide in about 2 months. BA.1 demonstrated considerable escape from neutralization by monoclonal antibodies (mAbs) and sera from vaccinated individuals (Cameroni et al., 2022; Cao et al., 2022; Carreño et al., 2022; Cele et al., 2022; Garcia-Beltran et al., 2022; Liu et al., 2022; Planas et al., 2021a; VanBlargan et al., 2022; Zost et al., 2020). BA.2 has then sharply increased in frequency in many countries, suggesting that it is more transmissible and possesses a selective advantage over BA.1. As of February 2022, BA.2 prevailed in many countries, including Denmark, the Philippines, South Africa and Belgium. BA.1 and BA.2 share numerous mutations in common, but 20 mutations in the spike differentiate the two sub-lineages (Fig. 1a). The sensitivity of BA.2 to neutralizing antibodies starts to be unveiled. Recent preprints and articles, using mainly pseudoviruses, indicated a drop in the neutralizing activity of sera from vaccine recipients against BA.2, relative to ancestral strains, to an extent comparable to BA.1 (Iketani et al., 2022; Yamasoba et al., 2022; Yu et al., 2022; Zhou et al., 2022). BA.2 also displays a marked decreased sensitivity to many neutralizing monoclonal antibodies (mAbs), when compared to previous strains (Cathcart et al., 2022; Iketani et al., 2022; Mykytyn et al., 2022; Yamasoba et al., 2022; Zhou et al., 2022).

**Figure 1:**
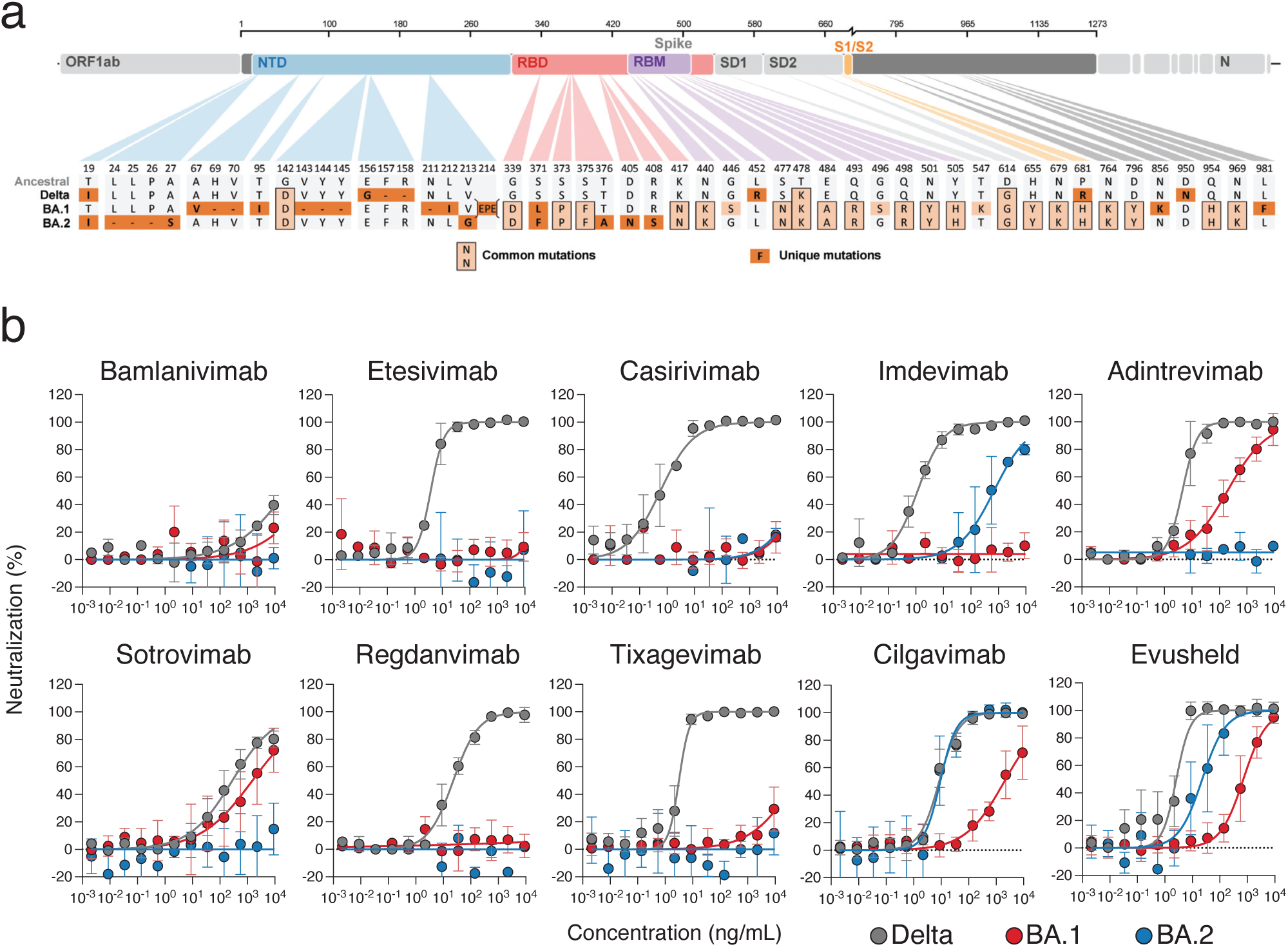
Sensitivity of Omicron BA.1 and BA.2 to therapeutic mAbs. **a**. Mutational landscape of Omicron BA.1 and BA.2 spike proteins. Domains of the protein are color-coded: NTD (N-Terminal Domain), Receptor-Binding Domain (RBD), Receptor-Binding Motif (RBM), subdomains 1 and 2 (SD1, SD2), region proximal to the furin cleavage site (S/S2). Mutations in the amino acid sequence are indicated in comparison to the ancestral Wuhan-Hu-1 sequence (GenBank: NC_045512). Light orange boxes indicate mutations shared by BA.1 and BA.2 and orange boxes indicates mutations unique to BA.1 and BA.2. **b**. Neutralization curves of monoclonal antibodies. Dose–response analysis of the neutralization by the indicated antibodies and by Evusheld, a combination of Cilgavimab and Tixagevimab). Data are mean ± s.d. of at least 2 independent experiments. The IC50 values for each antibody are presented in Extended Data Table 1.

Neutralizing mAbs targeting the receptor binding domain (RBD) of the SARS-CoV-2 spike, isolated from COVID-19 convalescent individuals demonstrated efficacy in preventing or treating disease in animal models and in humans (Crowe Jr., 2022). Some mAbs are used in combination, such as Ronapreve (Imdevimab and Etesivimab) from Regeneron and Evusheld (Cilgavimab and Tixagevimab) from AstraZeneca. Evusheld mAbs are modified in their Fc regions to improve half-life and decrease Fc-effector functions (Zost et al., 2020). Post-exposure administration of Ronapreve prevented 84% of infections in a randomized clinical trial, performed before Omicron circulation (O’Brien et al., 2021). In a preclinical model, Evusheld protected macaques from infection with an ancestral SARS-CoV-2 (Loo et al., 2022). A press release from AstraZeneca indicated that intra-muscular administration of Evusheld (300 mg) reduced symptomatic disease by 83% (AstraZeneca, 2021). The efficacy of Evusheld in preventing virus infection is not known. Both Ronapreve and Evushled received emergency use approval for pre-exposure prophylaxis (PrEP) in many countries. However, in cell culture systems, BA.1 is resistant to Casirivimab and Imdevimab, and partially evades Cilgavimab and Tixagevimab (Cameroni et al., 2022; Cao et al., 2022; Liu et al., 2022; Planas et al., 2021a). Different studies reported a 11- to 183-fold increase in the inhibitory concentration 50% (IC50) of Evusheld against BA.1 relative to ancestral strains (NIH, 2022). As BA.1 was becoming predominant, these results motivated the switch of emergency use from Ronapreve to Evusheld for PreP in immunocompromised individuals. Besides Ronapreve and Evusheld, other mAbs are in clinical use. For instance, Sotrovimab, a pan-coronavirus antibody is indicated for treatment of infected individuals at risk for severe disease (Agarwal et al., 2020). The relative capacity of mAbs to neutralize Omicron BA.1 and BA.2 sublineages is poorly characterized, with discordant preliminary results regarding mAbs such as Sotrovimab and Imdevimab. The clinical significance of the reduced sensitivity of Omicron BA.1 and BA.2 to neutralizing antibodies in cell culture remains unknown.

**Table 1:** IC50 of therapeutic monoclonal antibodies against Delta, Omicron BA.1 and BA.2

|  | Delta | BA.1 | BA.2 |
| --- | --- | --- | --- |
| Bamlanivimab | >9000 | >9000 | >9000 |
| Etesivimab | 3.8 | >9000 | >9000 |
| Casirivimab | 0.58 | >9000 | >9000 |
| Imdevimab | 1.2 | >9000 | 693 |
| Adintrevimab | 4.5 | 198 | >9000 |
| Regdavimab | 23 | >9000 | >9000 |
| Sotrovimab | 280 | 1508 | >9000 |
| Tixagevimab | 3.2 | >9000 | >9000 |
| Cilgavimab | 8.5 | 1988 | 9.3 |
| Evusheld | 2.6 | 715 | 23 |

IC50 (ng/mL)

## Results & discussion

We isolated a BA.2 variant from a nasopharyngeal swab which was initially sequenced at the National Reference Center of UZ/KU Leuven (Belgium). The virus was amplified by two passages on Vero E6 cells and re-sequenced (Pango lineage BA.2, 21L (Omicron) according to Nextstrain, GISAID accession ID: (EPI_ISL_10654979) (Fig. 1a). When compared to Delta, the BA.2 spike protein contained 28 changes, with 18 modifications that are shared with BA.1 (Fig. 1a). The modifications are dispersed throughout the spike but display a preferential accumulation in the N-terminal domain (NTD) and the RBD (Fig. 1a). Viral stocks were titrated using S-Fuse cells. These reporter cells become GFP+ upon infection, allowing rapid measurement of viral infectivity and neutralizing antibody activity (Buchrieser et al., 2020; Planas et al., 2021b). Syncytia were observed in BA.2-infected S-Fuse cells, with a size similar to those induced by BA.1 (not shown). Future experiments are warranted to determine affinity to ACE2 and other characteristics of the BA.2 spike.

We first measured the sensitivity of BA.2 to a panel of 9 mAbs that were or are currently in clinical use (Hansen et al., 2020; Jones et al., 2021; Kim et al., 2021; Pinto et al., 2020; Rappazzo et al., 2021; Shi et al., 2020; Zost et al., 2020). These mAbs belong to the 4 main classes of anti-RBD antibodies, depending on their binding site (Barnes et al., 2020; Liu et al., 2020; Taylor et al., 2021). In addition to the antibodies present in Ronapreve and Evusheld, we tested the following antibodies: Bamlanivimab and Etesevimab (class 2 and class 1, respectively) initially mixed in the Lilly cocktail and that are no longer in clinical use. Regdanvimab (Regkirona™) (Celltrion) is a class 1 antibody. Sotrovimab (Xevudy™) by GlaxoSmithKline and Vir Biotechnology is a class 3 antibody that targets an epitope outside of the receptor binding motif. Adintrevimab (ADG20, Adagio) binds to an epitope located in between class 1 and class 4 sites. We compared the activity of the 9 mAbs against Delta, Omicron BA.1 and BA.2 strains (Fig. 1b).

Seven antibodies (Bamlanivimab, Etesevimab, Casirivimab, Sotrovimab, Adintrevimab, Regdanvimab and Tixagevimab) were inactive against BA.2. The two other antibodies (Imdevimab and Cilgavimab) displayed an IC50 of 693 and 9 ng/ml, against BA.2, respectively (Fig. 1b and Table 1), meaning that they were more active against BA.2 than BA.1. The addition of Tixagevimab to Cilgavimab in the Evusheld cocktail was not more efficient than Cilgavimab alone (Fig. 1b and Table 1). These results are in line with recent reports (Iketani et al., 2022b; Yamasoba et al., 2022; Zhou et al., 2022) and highlight significant differences in the neutralization profile of BA.1 and BA.2.

We next measured antibody levels and neutralization activity in the sera of 29 immunocompromised individuals before and after administration of Evusheld (Table 2). Some subjects were previously treated with Ronapreve (n=18 out of 29). The first group of patients was a cohort of 8 individuals (6 females and 2 males) from the Centre Hospitalier Regional of Orléans, France. Pre-existing conditions were rheumatic arthritis (RA, n=5), kidney transplantation (n=2) and myelodysplasia (n=1). Most patients were treated with anti-CD20 (Rituximab) (n=5) and prednisone (n=4). They were previously vaccinated with three doses of BNT162b2 (Pfzier/BioNTech) and 3 had a 4^th^ dose. Three patients received Ronapreve as PrEP 4 to 7 weeks before Evusheld. The second group of 21 patients (13 females and 8 males) came from Hôpital Cochin in Paris. They were suffering from auto-immune diseases, including RA (n=2), vasculitis (n=17), polychondritis (n=1) and lupus (n=1). They were vaccinated with three doses of BNT162b2, except one who received two doses of ChadOX-1 (AstraZeneca) and one dose of mRNA-1273 (Moderna). Three patients received a 4^th^ dose of BNT162b2 and another had an history of COVID-19. They were mostly treated with Rituximab (n = 17). Fifteen out of the 21 individuals were already having PrEP with Ronapreve. None of the 29 subjects elicited antibodies above 264 Binding Antibody Unit (BAU)/mL after vaccination and were thus eligible to Evusheld PreP, according to the French health authority guidelines (HAS, 2021).

**Table 2:**
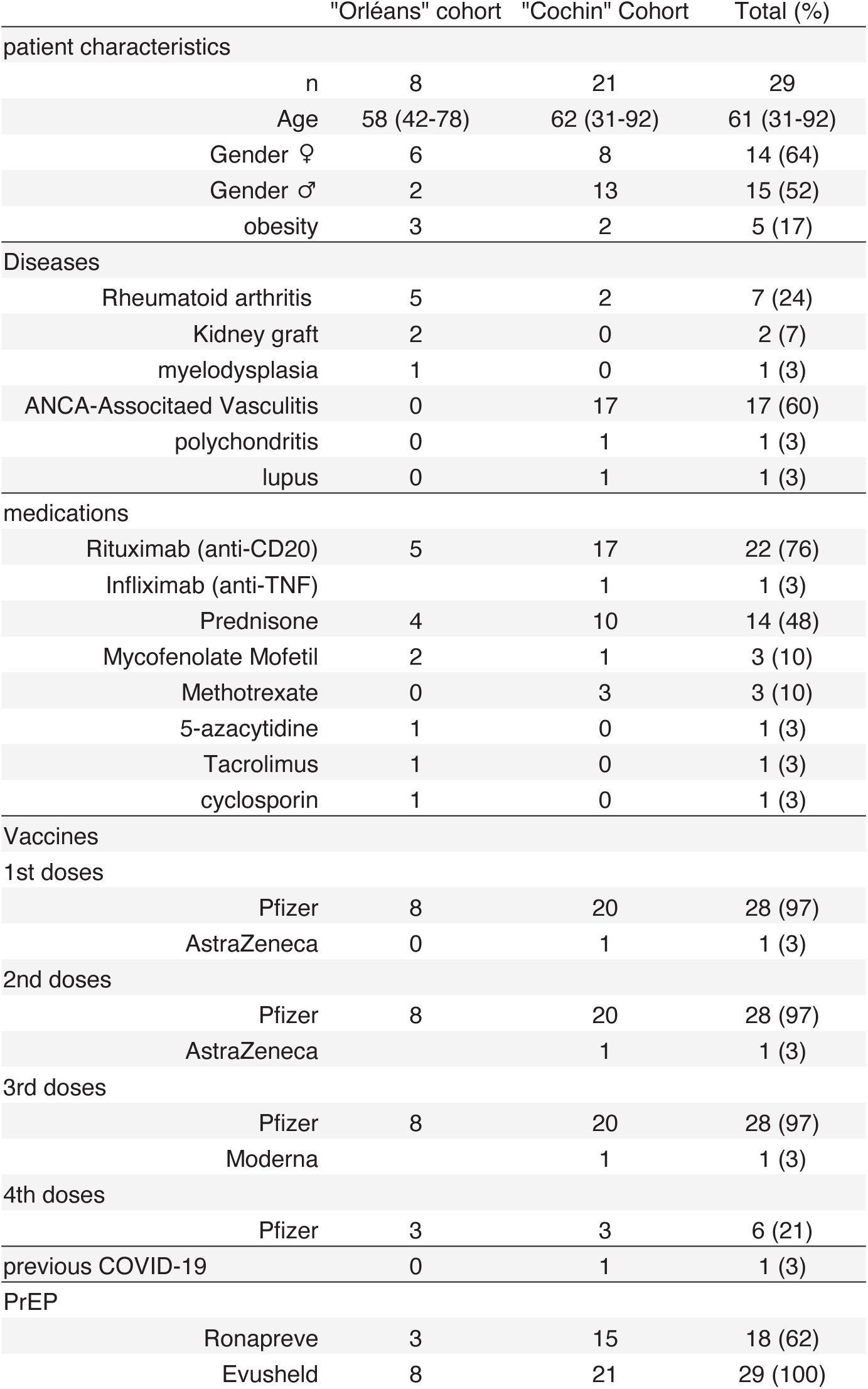
Characteristics of patients

|  | "Orléans" cohort | "Cochin" Cohort | Total (%) |
| --- | --- | --- | --- |
| patient characteristics |  |  |  |
| n | 8 | 21 | 29 |
| Age | 58 (42-78) | 62 (31-92) | 61 (31-92) |
| Gender ♀ | 6 | 8 | 14 (64) |
| Gender ♂ | 2 | 13 | 15 (52) |
| obesity | 3 | 2 | 5 (17) |
| Diseases |  |  |  |
| Rheumatoid arthritis | 5 | 2 | 7 (24) |
| Kidney graft | 2 | 0 | 2 (7) |
| myelodysplasia | 1 | 0 | 1 (3) |
| ANCA-Associated Vasculitis | 0 | 17 | 17 (60) |
| polychondritis | 0 | 1 | 1 (3) |
| lupus | 0 | 1 | 1 (3) |
| medications |  |  |  |
| Rituximab (anti-CD20) | 5 | 17 | 22 (76) |
| Infliximab (anti-TNF) |  | 1 | 1 (3) |
| Prednisone | 4 | 10 | 14 (48) |
| Mycophenolate Mofetil | 2 | 1 | 3 (10) |
| Methotrexate | 0 | 3 | 3 (10) |
| 5-azacytidine | 1 | 0 | 1 (3) |
| Tacrolimus | 1 | 0 | 1 (3) |
| cyclosporin | 1 | 0 | 1 (3) |
| Vaccines |  |  |  |
| 1st doses |  |  |  |
| Pfizer | 8 | 20 | 28 (97) |
| AstraZeneca | 0 | 1 | 1 (3) |
| 2nd doses |  |  |  |
| Pfizer | 8 | 20 | 28 (97) |
| AstraZeneca |  | 1 | 1 (3) |
| 3rd doses |  |  |  |
| Pfizer | 8 | 20 | 28 (97) |
| Moderna |  | 1 | 1 (3) |
| 4th doses |  |  |  |
| Pfizer | 3 | 3 | 6 (21) |
| previous COVID-19 | 0 | 1 | 1 (3) |
| PrEP |  |  |  |
| Ronapreve | 3 | 15 | 18 (62) |
| Evusheld | 8 | 21 | 29 (100) |

**Table 3:** Summary of breakthrough cases

| case | diagnostic | strain | days post<br>Evusheld | anti-S<br>(BAU/mL) | Neutralization<br>BA.1 (ED50) | COVID-19 |
| --- | --- | --- | --- | --- | --- | --- |
| 1 | PCR +<br>screening | Omicron | 15 | 9,630 | 351 | Mild |
| 2 | PCR +<br>screening | Omicron | 12 | 5,736 | 7,5 | Mild |
| 3 | PCR +<br>screening | Omicron | 21 | 1,786 | 36 | Mild |
| 4 | PCR +<br>sequencing | BA.1 | 23 | 4,536 | 31 | Severe |

We first analyzed the 8 individuals from the Orléans cohort, as longitudinal samples were available (Fig. 2a). We used the S-Flow assay to quantify anti-spike IgGs in sera collected at days 0, 3, 15 and 30 post-Evusheld administration. Day 30 sampling was only available for 4 individuals. In the 5 Ronapreve-naïve individuals, administration of Evusheld lead to a sharp increase of anti-spike IgGs (from 5-57 BAU/mL before treatment to 195-1290 BAU/mL after treatment) (Fig. 2a). As expected, the three individuals who initially received Ronapreve had anti-spike antibodies (788-1016 BAU/mL) at the time of Evusheld administration (day 0), with no detectable impact of Evusheld on antibody levels (Fig. 2a). In all patients, levels of anti-spike antibodies were stable or slightly increasing between days 3 and 30 (Fig. 2a).

**Figure 2:**
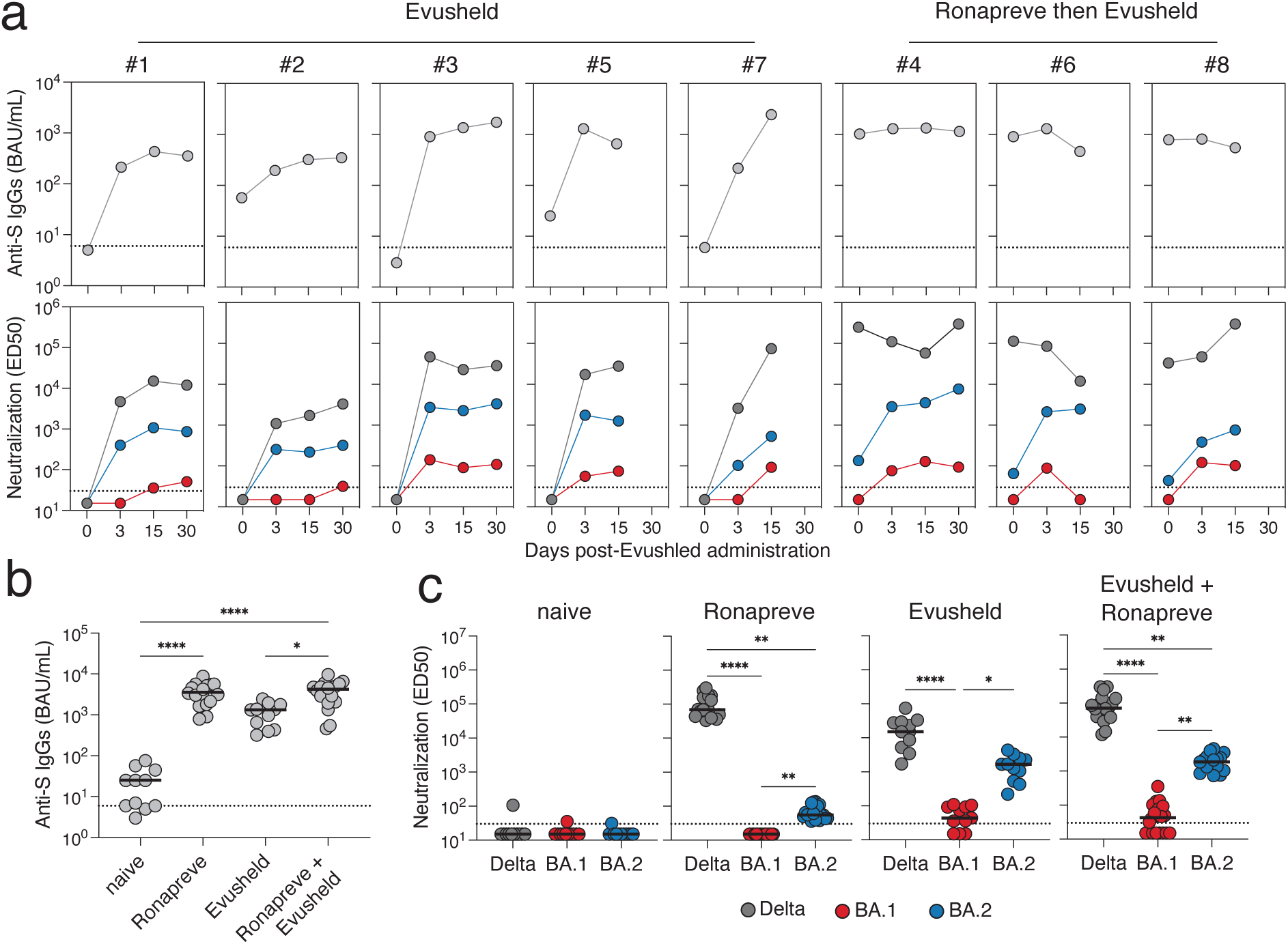
Neutralization of Delta and Omicron BA.1 and BA.2 by sera of immunocompromised individuals receiving Ronapreve and/or Evusheld as a pre-exposure prophylaxis. **a**. Eight individuals from the Orléans Cohort were followed longitudinally, prior and after Evusheld administration. Anti-S IgGs were measured using the flow cytometry-based S-Flow assay (top panel). Neutralization of Delta, Omicron BA.1 and BA.2 was measured with the S-fuse assay (bottom panel). The dotted lines indicate the limit of detection of the assays. Three individuals received first Ronapreve and then Evusheld. **b**. Anti-S IgGs levels in sera of individuals prior to PrEP (naïve; n=11), treated with Ronapreve (n=18), Evusheld (n=11), or with both Ronapreve and Evuslehd (n=18). **c**. Sero-neutralisation of Delta, Omicron BA.1 and BA.2 in the same individuals as in b. Two-sided Friedman tests with Dunn’s multiple-comparison correction was performed to compare the different groups; *P < 0.05, **P < 0.01, ***P < 0.001.

We then measured the neutralizing activity of the sera against Delta, Omicron BA.1 and BA.2 by calculating Effective Dilution 50% titers (ED50) with the S-Fuse assay (Fig. 2a). None of the 5 Ronapreve-naïve individuals had detectable neutralization activity at day 0. Evusheld administration led to a sharp increase of neutralizing activity against Delta, with ED50s between 788 and 1016. For the three individuals having previously received Ronapreve, Evusheld administration did not increase their levels of neutralization against Delta. In line with in vitro experiments (Fig. 1b and (Cao et al., 2022; Planas et al., 2021a), neither Ronapreve-naïve nor Ronapreve-treated individuals neutralized BA.1. After Evusheld treatement, 7 out of 8 individuals neutralized BA.1 at different time points between days 3 and 30. Titers were however very low, ranging from 27 to 128 at day 15. A low level of BA.2 neutralization was detectable in the 3 Ronapreve-treated individuals, in line with the ability of Imdevimab to neutralize BA.2 (Fig. 1b). The 5 Ronapreve-naïve patients did not neutralize BA.2 at day 0. Evusheld administration raised BA.2 neutralization in all patients, with titers reaching up to an ED50 of 3534 at day 15 (Fig. 2b). Neutralization titers for the three viral lineages were stable for 6 out of 8 patients, consistently with the Evusheld long half-life (Loo et al., 2022).

We extended this analysis to the 21 individuals of the second group, who were sampled at a single time point, 15 to 30 days after Evusheld administration. We mixed the results obtained with the first group of 8 individuals at day 15, to analyze together the 29 individuals. The 9 Ronapreve-naïve individuals had low levels of anti-spike antibodies (below 264 BAU/mL), reflecting the inefficacy of the vaccination (Fig. 2b). Ronapreve or Evusheld therapy strongly and similarly increased anti-spike IgGs in the sera (median of 3263 BAU/mL and 1321 BAU/mL) (Fig. 2b). These levels were not higher in individuals that successively received the two treatments (Fig. 2b). We next measured neutralization titers in the 29 sera (Fig. 2c). The untreated individuals did not neutralize any of the three strains. Ronapreve-treated individuals efficiently neutralized Delta, were inactive against BA.1 and poorly neutralized BA.2. Sera from Evusheld and Ronapreve+Evusheld treated individuals were efficient against Delta, barely neutralized BA.1 (ED50 of 44 and 42, respectively) and quite efficiently neutralized BA.2 (ED50 of 1673 and 1882, representing a 38- and 49-fold decrease compared to Delta) (Fig. 2c). After Evusheld administration, 8 out of 11 individuals, who did not previously receive Ronapreve had neutralization activity against BA.1 in their sera, and all neutralized BA.2. This confirmed that Evusehld is more active against BA.2 than BA.1. There was no major difference in the neutralization titers in individuals having received only Evusheld or the successive combination of Ronapreve and Evusheld (Fig. 2c). The neutralizing activity against Delta correlated to anti-spike IgG levels, whereas this was not the case for BA.1 and BA.2 (Fig. Sup. 1). This reflects an uncoupling of the capacity of the antibodies to bind to the spike from the ancestral Wuhan strain, and to neutralize Omicron BA.1 and BA.2 strains. Altogether, these data show that administration of Evusheld in immunocompromised individuals renders their sera able to poorly neutralize BA.1 and to act more efficiently on BA.2.

In agreement with a decreased sero-neutralization activity of Evusheld-treated individuals against BA.1, we observed 4 breakthrough infections among the 29 participants. A summary of the cases is provided Table 2 and Supplementary Fig. 2, along with the serology and neutralization data of the closest sampling point. A PCR screening confirmed Omicron infection for the 4 cases but did not allow distinction between BA.1 and BA.2. However, at the time of the sampling, BA.1 represented 90% of sequenced cases whereas BA.2 was detected in <10% of cases in France. A sequence was performed only for case #4 and confirmed BA.1 infection. Three out of the 4 patients received Sotrovimab after diagnosis, according to French guidelines. Three cases were classified as a mild disease, whereas case #4 was severe and required hospitalization. Despite detection of high levels of anti-spike antibodies in the sera, the neutralization titers against BA.1 were low and ranged between <7.5 and 351 for the 4 individuals (Table 2 and Supplementary Fig. 2). These four cases indicate that Evusheld does neither protect against Omicron infection nor fully prevent against severe disease.

We highlight here significant differences not only between Delta and Omicron, but also between BA.1 and BA.2 Omicron sublineages with regards to their sensitivity to therapeutic mAbs. Considering that these variants have sequentially dominated the pandemic in a few months, and the potentially critical impact of these results on treatment outcome of immunocompromised patients, our results support the utility of performant genomic surveillance throughout the pandemic. If these results had to translate into altered treatment efficacy during clinical trials, a rapid genotyping will need to be introduced in clinical practice in order to inform for the more adapted treatment. Our results also show that measuring antibody levels with standard serology assays that currently use an ancestral spike antigen, without adapted neutralization tests, cannot be used as a marker of clinical efficacy.

Our study has limitations. The relatively low number of individuals analyzed did not allow evaluating the clinical efficacy of Evusheld against BA.2. We did not have access to nasopharyngeal samples of the patients. Measuring antibody levels and neutralization in these swabs will help providing insights into the capacity of mAbs to neutralize Omicron sub-lineages at the infection site. We did not test BA.1.1 and BA.3 sub-lineages of Omicron. Future experiments with these viruses are needed to determine the antiviral activity of mAbs against the full landscape of the Omicron clade, which we recently proposed to be considered as a distinct SARS-CoV-2 serotype from ancestral strains and previous variants (Simon-Loriere and Schwartz, 2022).

Based on our observation of breakthrough infections and awaiting clinical trials which will provide a complete evaluation of the impact of BA.2 on the treatment efficacy of mAbs, we expect more frequent treatment failures. It is also possible that the progressive accumulation of further mutations will increase the level of resistance of BA.1 or BA.2 to mAbs during prolonged infection. The low or intermediate sensitivity to Ronapreve and Evusheld, when used as a pre-exposure prophylaxis in immunocompromised persons at risk for severe disease is of potential concern. The risk that further escape mutations will arise in these patients is higher compared to Delta. We therefore recommend a close follow-up of these patients, particularly in case of prolonged infection despite treatment.

## Methods

### Design

No statistical methods were used to predetermine sample size. The experiments were not randomized and the investigators were not blinded to allocation during experiments and outcome assessment. Our research complies with all relevant ethical regulation.

### Cohorts

Immunocompromised individuals receiving Evusheld were recruited in two centers (CHR d’Orléans and Hôpital Cochin), in the French cities of Orléans and Paris. The “Orléans” cohort is an ongoing prospective, monocentric, longitudinal, observational cohort clinical study aiming to describe the kinetic of neutralizing antibodies after SARS-CoV-2 infection or vaccination (ClinicalTrials.gov Identifier: NCT04750720). This study was approved by the Est II (Besançon) ethical committee. At enrolment, written informed consent was collected and participants completed a questionnaire which covered sociodemographic characteristics, clinical information, and data related to anti-SARS-CoV-2 vaccination. Blood sampling was performed the day of Evusheld infusion and after 3 days, 15 days and 1 month. The “Cochin” cohort is a prospective, monocentric, longitudinal, observational clinical study (NCT04870411) enrolling immunocompromised individuals with rheumatic diseases, aiming at describing immunological responses to COVID-19 vaccine in patients with autoimmune and inflammatory diseases treated with immunosuppressants and/or biologics. Ethics approval was obtained by Comite de Protection des Personnes Nord-Ouest II. Leftover sera from usual care were used from these individuals in the setting of the local biological samples collection (RAPIDEM). A written informed consent was collected for all participants. None of the study participants received compensation.

### Viral strains

The Delta strain was isolated from a nasopharyngeal swab of a hospitalized patient returning from India (Planas et al., 2021c). The swab was provided and sequenced by the laboratory of Virology of Hopital Européen Georges Pompidou (Assistance Publique – Hopitaux de Paris). The Omicron strain was supplied and sequenced by the NRC UZ/KU Leuven (Leuven, Belgium) (Planas et al., 2021a). The BA.2 strain was isolated from a nasopharyngeal swab sampled in January 2022 from a 5-10 years old male patient. The sample was sequenced in the context of active surveillance by the NRC UZ/KU Leuven (Leuven, Belgium), showing an average coverage of 989x for the Omicron BA.2 genome, after which it was cultured on Vero E6 cells. We noted an additional mutation in the spike of our BA.2 isolate (R682W) compared to the primary sample from which it was isolated, although this mutation was already present at low frequency in the original swab. Both patients provided informed consent for the use of the biological materials. The sequences of the isolates were deposited on GISAID immediately after their generation, with the following Delta ID: EPI_ISL_2029113; Omicron ID: EPI_ISL_6794907. Omicron BA.2 GISAID ID: EPI_ISL_10654979. Titration of viral stocks was performed on Vero E6, with a limiting dilution technique allowing a calculation of TCID50, or on S-Fuse cells.

### Monoclonal antibodies

Bamlanivimab, Casirivimab, Etesevimab, Imdevimab, Cilgavimab, Tixagevimab and Sotrovimab were provided by CHR Orleans. Adintrevimab (ADG20) and Regdanvimab (CT-P59), were produced as previously described (Planas et al., 2021d).

### S-Fuse neutralization assay

U2OS-ACE2 GFP1-10 or GFP 11 cells, also termed S-Fuse cells, become GFP+ when they are productively infected by SARS-CoV-2. Cells tested negative for mycoplasma. Cells were mixed (ratio 1:1) and plated at 8×103 per well in a μClear 96-well plate (Greiner Bio-One). The indicated SARS-CoV-2 strains were incubated with serially diluted mAb or sera for 15 minutes at room temperature and added to S-Fuse cells. The sera were heat-inactivated 30 min at 56 °C before use. 18 hours later, cells were fixed with 2% PFA, washed and stained with Hoechst (dilution 1:1,000, Invitrogen). Images were acquired with an Opera Phenix high content confocal microscope (PerkinElmer). The GFP area and the number of nuclei were quantified using the Harmony software (PerkinElmer). The percentage of neutralization was calculated using the number of syncytia as value with the following formula: 100 x (1 – (value with serum – value in “non-infected”)/(value in “no serum” – value in “non-infected”)). Neutralizing activity of each serum was expressed as the half maximal effective dilution (ED50). ED50 values (in μg/ml for mAbs and in dilution values for sera) were calculated with a reconstructed curve using the percentage of the neutralization at the different concentrations.

### Anti-spike serology

The S-Flow assay uses 293T cells stably expressing the spike protein (293T Spike cells) and 293T control cells as control to detect anti-S antibodies by flow cytometry (Grzelak et al., 2021).. Briefly, the cells were incubated at 4°C for 30 min with sera (1:300 dilution) in PBS containing 0.5% BSA and 2 mM EDTA. Cells were then washed with PBS and stained with an anti-human IgG Fc Alexafluor 647 antibody (109-605-170, Jackson ImmunoResearch). After 30min at 4°C, cells were washed with PBS and fixed for 10 min using 4% paraformaldehyde (PFA). A standard curve with serial dilutions of a human anti-spike monoclonal antibody (mAb48) was acquired in each assay to standardize the results. Data were acquired on an Attune NxT instrument (Life Technologies) analyzed with FlowJo v.10 software (TriStar). The sensitivity is 99.2% with a 95% confidence interval of 97.69–99.78% and the specificity is 100% (98.5–100%)(Grzelak et al., 2021). To determine BAU/mL, we analyzed a series of vaccinated (n=144), convalescent (n=59) samples and WHO international reference sera (20/136 and 20/130) on S-Flow and on two commercially available ELISA (Abbott 147 and Beckmann 56). Using this dataset, we performed a Passing-Pablok regression, which shows that the relationship between BU and BAU/mL is linear, allowing calculation of BAU/mL using S-Flow data (Hadjadj et al., 2022).

### Statistical analysis

Flow cytometry data were analyzed with FlowJo v10 software (TriStar). Calculations were performed using Excel 365 (Microsoft). Figures were drawn on Prism 9 (GraphPad Software). Statistical analysis was conducted using GraphPad Prism 9. Statistical significance between different groups was calculated using the tests indicated in each figure legend.

### Data availability

All data supporting the findings of this study are available within the article or from the corresponding authors upon request. Source data are provided with this paper.

## Data Availability

All data supporting the findings of this study are available within the article or from the corresponding authors upon request.

## Acknowledgments

We thank the European Health Emergency Preparedness and Response Authority, (HERA) for supporting the work being done at Institut Pasteur and UK Leuven. We thank Julian Buchrieser, Julien Puech and Fabiana Gambaro for their help with the sequencing data analysis. We thank patients who participated to this study, members of the Virus and Immunity Unit and other teams for discussions and help, Nathalie Aulner and the UtechS Photonic BioImaging (UPBI) core facility (Institut Pasteur), a member of the France BioImaging network, for image acquisition and analysis. The Opera system was co-funded by Institut Pasteur and the Région ile de France (DIM1Health). We thank the KU Leuven University authorities and Jef Arnout, Bruno Lambrecht, Chris Van Geet and Luc Sels for their support. We thank Fabienne Peira, Vanessa Legros and Laura Courtellemont for their help with the cohorts. UZ Leuven, as national reference center for respiratory pathogens, is supported by Sciensano, which is gratefully acknowledged.

## Figures Sup and tables

**Supplementary Figure 1:**
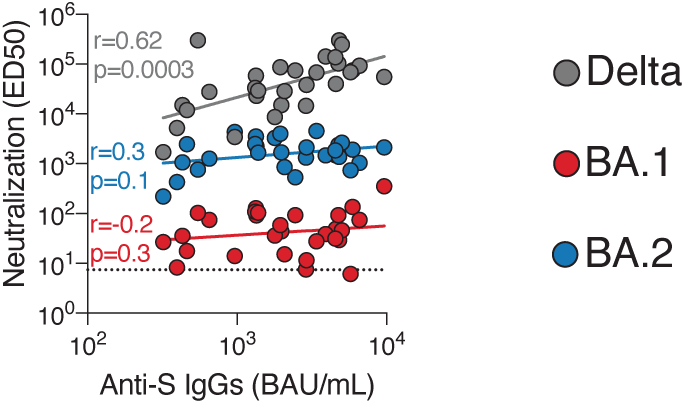
Correlation of neutralization capacity and anti-S antibody levels in individuals having received Ronapreve and/or Evusheld. Spearman non-parametric correlations of neutralizing antibody titers against Delta, Omicron BA.1 and BA.2 and the level of anti-S IgG. R and p-values are indicated.

**Supplementary Figure 2:**
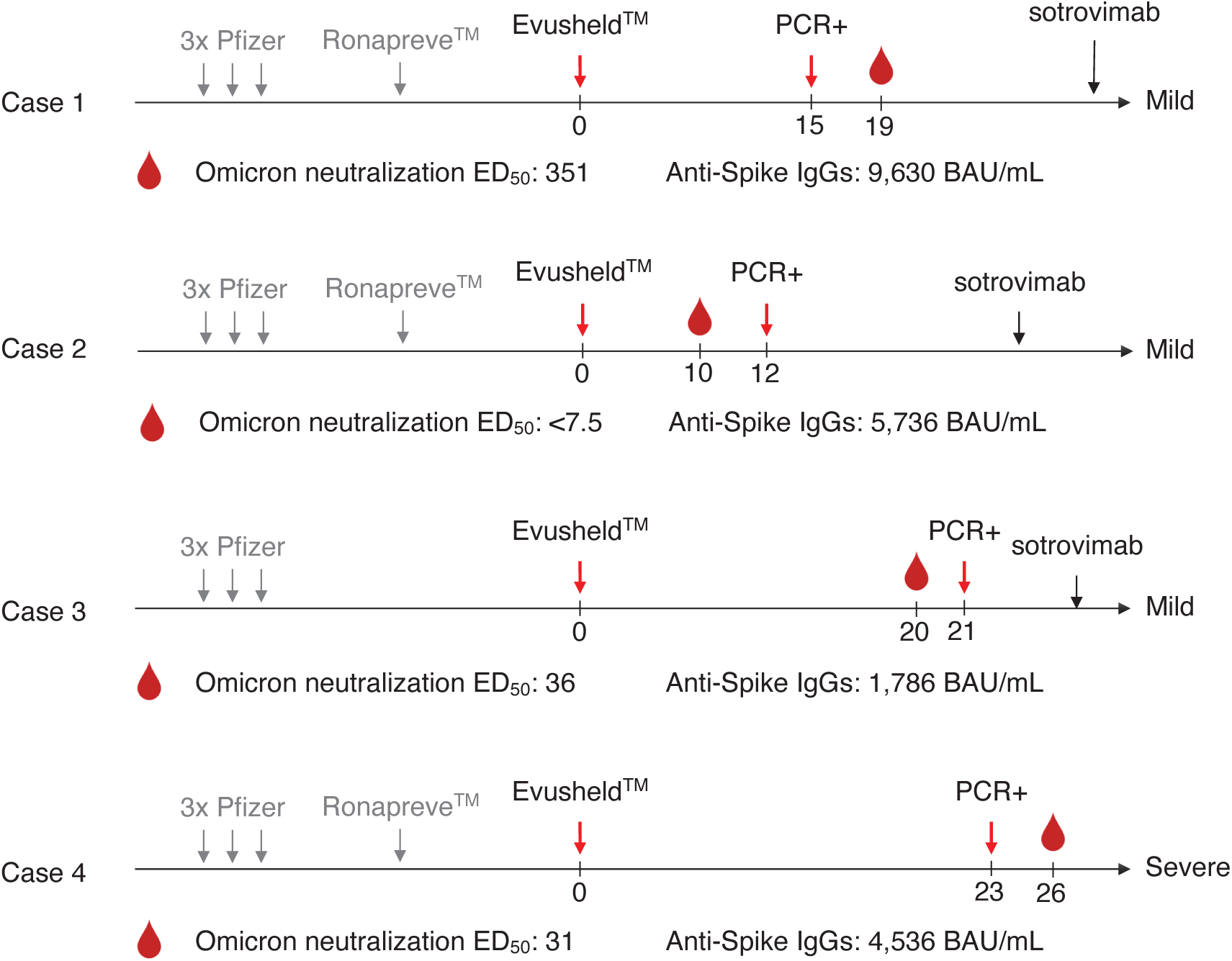
Report of four Omicron breakthrough infections in Evusheld treated patients. A timeline indicates the key events for each of the 4 Omicron breakthrough cases. Patients’ characteristics and antibody measurement of the closest sampling point are indicated.

**Supplementary Table 1: IC50 of therapeutic monoclonal antibodies against Delta and Omicron BA1 and BA2**.

**Supplementary Table 2: Characteristics of patients**.

**Supplementary Table 3: Summary of breakthrough cases**.

## References

Agarwal, A., Rochwerg, B., Lamontagne, F., Siemieniuk, R.A., Agoritsas, T., Askie, L., Lytvyn, L., Leo, Y.-S., Macdonald, H., Zeng, L., et al. (2020). A living WHO guideline on drugs for covid-19. Bmj 370, m3379.

AstraZeneca (2021). New analyses of two AZD7442 COVID-19 Phase III trials in high-risk populations confirm robust efficacy and long-term prevention. https://www.astrazeneca.com/media-centre/press-releases/2021/new-analyses-of-two-azd7442-covid-19-phase-iii-trials-in-high-risk-populations-confirm-robust-efficacy-and-long-term-prevention.html

Barnes, C.O., Jette, C.A., Abernathy, M.E., Dam, K.-M.A., Esswein, S.R., Gristick, H.B., Malyutin, A.G., Sharaf, N.G., Huey-Tubman, K.E., Lee, Y.E., et al. (2020). SARS-CoV-2 neutralizing antibody structures inform therapeutic strategies. Nature 588, 682–687.

Buchrieser, J., Dufloo, J., Hubert, M., Monel, B., Planas, D., Rajah, M.M., Planchais, C., Porrot, F., Guivel-Benhassine, F., Werf, S.V. der, et al. (2020). Syncytia formation by SARS-CoV-2 infected cells. Embo J e2020106267.

Cameroni, E., Bowen, J.E., Rosen, L.E., Saliba, C., Zepeda, S.K., Culap, K., Pinto, D., VanBlargan, L.A., Marco, A.D., Iulio J. di, et al. (2022). Broadly neutralizing antibodies overcome SARS-CoV-2 Omicron antigenic shift. Nature 602, 664–670.

Cao, Y., Wang, J., Jian, F., Xiao, T., Song, W., Yisimayi, A., Huang, W., Li, Q., Wang, P., An, R., et al. (2022). Omicron escapes the majority of existing SARS-CoV-2 neutralizing antibodies. Nature 602, 657–663.

Carreño, J.M., Alshammary, H., Tcheou, J., Singh, G., Raskin, A.J., Kawabata, H., Sominsky, L.A., Clark, J.J., Adelsberg, D.C., Bielak, D.A., et al. (2022). Activity of convalescent and vaccine serum against SARS-CoV-2 Omicron. Nature 602, 682–688.

Cathcart, A.L., Havenar-Daughton, C., Lempp, F.A., Ma, D., Schmid, M.A., Agostini, M.L., Guarino, B., iulio, J.D., Rosen, L.E., Tucker, H., et al. (2022). The dual function monoclonal antibodies VIR-7831 and VIR-7832 demonstrate potent in vitro and in vivo activity against SARS-CoV-2. Biorxiv 2021.03.09.434607.

Cele, S., Jackson, L., Khoury, D.S., Khan, K., Moyo-Gwete, T., Tegally, H., San, J.E., Cromer, D., Scheepers, C., Amoako, D.G., et al. (2022). Omicron extensively but incompletely escapes Pfizer BNT162b2 neutralization. Nature 602, 654–656.

Garcia-Beltran, W.F., Denis, K.J.St., Hoelzemer, A., Lam, E.C., Nitido, A.D., Sheehan, M.L., Berrios, C., Ofoman, O., Chang, C.C., Hauser, B.M., et al. (2022). mRNA-based COVID-19 vaccine boosters induce neutralizing immunity against SARS-CoV-2 Omicron variant. Cell 185, 457–466.e4.

Grzelak, L., Velay, A., Madec, Y., Gallais, F., Staropoli, I., Schmidt-Mutter, C., Wendling, M.-J., Meyer, N., Planchais, C., Rey, D., et al. (2021). Sex Differences in the Evolution of Neutralizing Antibodies to Severe Acute Respiratory Syndrome Coronavirus 2. J Infect Dis 224, 983–988.

Hadjadj, J., Planas, D., Ouedrani, A., Buffier, S., Delage, L., Nguyen, Y., Bruel, T., Stolzenberg, M.-C., Staropoli, I., Ermak, N., et al. (2022). Immunogenicity of BNT162b2 vaccine against the Alpha and Delta variants in immunocompromised patients with systemic inflammatory diseases. Ann Rheum Dis annrheumdis-2021-221508.

Hansen, J., Baum, A., Pascal, K.E., Russo, V., Giordano, S., Wloga, E., Fulton, B.O., Yan, Y., Koon, K., Patel, K., et al. (2020). Studies in humanized mice and convalescent humans yield a SARS-CoV-2 antibody cocktail. Science 369, 1010–1014.

HAS (2021). EVUSHELD (tixagévimab/cilgavimab) DÉCISION D’ACCÈS PRÉCOCE. https://www.has-sante.fr/jcms/p_3304078/fr/decision-n-2021-0312/dc/sem-du-9-decembre-2021-du-college-de-la-haute-autorite-de-sante-portant-autorisation-d-acces-precoce-de-la-specialite-evusheld-tixagevimab/cilgavimab

Iketani, S., Liu, L., Guo, Y., Liu, L., Chan, J.F.-W., Huang, Y., Wang, M., Luo, Y., Yu, J., Chu, H., et al. (2022). Antibody evasion properties of SARS-CoV-2 Omicron sublineages. Nature 1–1. https://doi.org/10.1038/s41586-022-04594-4

Jones, B.E., Brown-Augsburger, P.L., Corbett, K.S., Westendorf, K., Davies, J., Cujec, T.P., Wiethoff, C.M., Blackbourne, J.L., Heinz, B.A., Foster, D., et al. (2021). The neutralizing antibody, LY-CoV555, protects against SARS-CoV-2 infection in nonhuman primates. Sci Transl Med 13, eabf1906.

Crowe, Jr., J.E.C. (2022). Human Antibodies for Viral Infections. Annu Rev Immunol 40. 10.1146/annurev-immunol-042718-041309

Kim, C., Ryu, D.-K., Lee, J., Kim, Y.-I., Seo, J.-M., Kim, Y.-G., Jeong, J.-H., Kim, M., Kim, J.-I., Kim, P., et al. (2021). A therapeutic neutralizing antibody targeting receptor binding domain of SARS-CoV-2 spike protein. Nat Commun 12, 288.

Liu, L., Wang, P., Nair, M.S., Yu, J., Rapp, M., Wang, Q., Luo, Y., Chan, J.F.-W., Sahi, V., Figueroa, A., et al. (2020). Potent neutralizing antibodies against multiple epitopes on SARS-CoV-2 spike. Nature 584, 450–456.

Liu, L., Iketani, S., Guo, Y., Chan, J.F.-W., Wang, M., Liu, L., Luo, Y., Chu, H., Huang, Y., Nair, M.S., et al. (2022). Striking antibody evasion manifested by the Omicron variant of SARS-CoV-2. Nature 602, 676–681.

Loo, Y.-M., McTamney, P.M., Arends, R.H., Abram, M.E., Aksyuk, A.A., Diallo, S., Flores, D.J., Kelly, E.J., Ren, K., Roque, R., et al. (2022). The SARS-CoV-2 monoclonal antibody combination, AZD7442, is protective in non-human primates and has an extended half-life in humans. Sci Transl Med eabl8124.

Mykytyn, A.Z., Rissmann, M., Kok, A., Rosu, M.E., Schipper, D., Breugem, T.I., Doel P.B. van den, Chandler, F., Bestebroer, T., Wit M. de, et al. (2022). Omicron BA.1 and BA.2 are antigenically distinct SARS-CoV-2 variants. Biorxiv 2022.02.23.481644.

NIH (2022). All Variants | Reported in vitro Therapeutic Activity. https://opendata.ncats.nih.gov/variant/summary

O’Brien, M.P., Forleo-Neto, E., Musser, B.J., Isa, F., Chan, K.-C., Sarkar, N., Bar, K.J., Barnabas, R.V., Barouch, D.H., Cohen, M.S., et al. (2021). Subcutaneous REGEN-COV Antibody Combination to Prevent Covid-19. New Engl J Med 385, 1184–1195.

Pinto, D., Park, Y.-J., Beltramello, M., Walls, A.C., Tortorici, M.A., Bianchi, S., Jaconi, S., Culap, K., Zatta, F., Marco, A.D., et al. (2020). Cross-neutralization of SARS-CoV-2 by a human monoclonal SARS-CoV antibody. Nature 583, 290–295.

Planas, D., Saunders, N., Maes, P., Guivel-Benhassine, F., Planchais, C., Buchrieser, J., Bolland, W.-H., Porrot, F., Staropoli, I., Lemoine, F., et al. (2021a). Considerable escape of SARS-CoV-2 Omicron to antibody neutralization. Nature 1–7.

Planas, D., Bruel, T., Grzelak, L., Guivel-Benhassine, F., Staropoli, I., Porrot, F., Planchais, C., Buchrieser, J., Rajah, M.M., Bishop, E., et al. (2021b). Sensitivity of infectious SARS-CoV-2 B.1.1.7 and B.1.351 variants to neutralizing antibodies. Nat Med 27, 917–924.

Planas, D., Veyer, D., Baidaliuk, A., Staropoli, I., Guivel-Benhassine, F., Rajah, M.M., Planchais, C., Porrot, F., Robillard, N., Puech, J., et al. (2021c). Reduced sensitivity of SARS-CoV-2 variant Delta to antibody neutralization. Nature 596, 276–280.

Planas, D., Saunders, N., Maes, P., Guivel-Benhassine, F., Planchais, C., Buchrieser, J., Bolland, W.-H., Porrot, F., Staropoli, I., Lemoine, F., et al. (2021d). Considerable escape of SARS-CoV-2 Omicron to antibody neutralization. Nature 1–7. https://doi.org/10.1038/d41586-021-03827-2

Rappazzo, C.G., Tse, L.V., Kaku, C.I., Wrapp, D., Sakharkar, M., Huang, D., Deveau, L.M., Yockachonis, T.J., Herbert, A.S., Battles, M.B., et al. (2021). Broad and potent activity against SARS-like viruses by an engineered human monoclonal antibody. Sci New York N Y 371, 823– 829.

Shi, R., Shan, C., Duan, X., Chen, Z., Liu, P., Song, J., Song, T., Bi, X., Han, C., Wu, L., et al. (2020). A human neutralizing antibody targets the receptor-binding site of SARS-CoV-2. Nature 584, 120–124.

Simon-Loriere, E., and Schwartz, O. (2022). Towards SARS-CoV-2 serotypes? Nat Rev Microbiol 1–2.

Taylor, P.C., Adams, A.C., Hufford, M.M., Torre, I. de la, Winthrop, K., and Gottlieb, R.L. (2021). Neutralizing monoclonal antibodies for treatment of COVID-19. Nat Rev Immunol 21, 382–393.

VanBlargan, L.A., Errico, J.M., Halfmann, P.J., Zost, S.J., Crowe, J.E., Purcell, L.A., Kawaoka, Y., Corti, D., Fremont, D.H., and Diamond, M.S. (2022). An infectious SARS-CoV-2 B.1.1.529 Omicron virus escapes neutralization by therapeutic monoclonal antibodies. Nat Med 1–6.

Viana, R., Moyo, S., Amoako, D.G., Tegally, H., Scheepers, C., Althaus, C.L., Anyaneji, U.J., Bester, P.A., Boni, M.F., Chand, M., et al. (2022). Rapid epidemic expansion of the SARS-CoV-2 Omicron variant in southern Africa. Nature 1–10. https://doi.org/10.1038/s41586-022-04411-y

Yamasoba, D., Kimura, I., Nasser, H., Morioka, Y., Nao, N., Ito, J., Uriu, K., Tsuda, M., Zahradnik, J., Shirakawa, K., et al. (2022). Virological characteristics of SARS-CoV-2 BA.2 variant. Biorxiv 2022.02.14.480335.

Yu, J., Collier, A.Y., Rowe, M., Mardas, F., Ventura, J.D., Wan, H., Miller, J., Powers, O., Chung, B., Siamatu, M., et al. (2022). Comparable Neutralization of the SARS-CoV-2 Omicron BA.1 and BA.2 Variants. Medrxiv 2022.02.06.22270533.

Zhou, H., Tada, T., Dcosta, B.M., and Landau, N.R. (2022). Neutralization of SARS-CoV-2 Omicron BA.2 by Therapeutic Monoclonal Antibodies. Biorxiv 2022.02.15.480166.

Zost, S.J., Gilchuk, P., Case, J.B., Binshtein, E., Chen, R.E., Nkolola, J.P., Schäfer, A., Reidy, J.X., Trivette, A., Nargi, R.S., et al. (2020). Potently neutralizing and protective human antibodies against SARS-CoV-2. Nature 584, 443–449.

